# Associations Between Reported Post-COVID-19 Symptoms and Subjective Well-Being, Israel, July 2021 -April 2022

**DOI:** 10.1101/2022.10.09.22280878

**Authors:** Yanay Gorelik, Amiel Dror, Hiba Zayyad, Ofir Wertheim, Kamal Abu Jabal, Saleh Nazzal, Paul Otiku, Jelte Elsinga, Daniel Glikman, Michael Edelstein

## Abstract

The impact of individual symptoms reported post-COVID-19 on subjective well-being (SWB) is unknown. We described associations between SWB and selected reported symptoms following SARS-CoV-2 infection. We analysed reported symptoms and subjective well being from 2295 participants (of which 576 reporting previous infection) in an ongoing longitudinal cohort study taking place in Israel. We estimated changes in SWB associated with reported selected symptoms at three follow-up time points (3-6, 6-12, and 12-18 months post infection) among participants reporting previous SARS-CoV-2 infection, adjusted for key demographic variables, using linear regression. Our results suggest that the biggest and most sustained changes in SWB stems from non-specific symptoms (fatigue −7.7 percentage points (pp), confusion/ lack of concentration −10.7 pp, and sleep disorders −11.5pp, p<0.005), whereas the effect of system-specific symptoms, such as musculoskeletal symptoms (weakness in muscles and muscle pain) on SWB, are less profound and more transient. Taking a similar approach for other symptoms and following individuals over time to describe trends in SWB changes attributable to specific symptoms will help understand the post-acute phase of COVID-19 and how it should be defined and better managed.

## Introduction

As of August 2022, over 585,000,000 cases of COVID-19 had been reported globally, including more than 4,600,000 cases in Israel, approximately 48% of the population.[1] As a result of suboptimal surveillance and lack of testing, true figures are likely to be much higher [2]. Evidence suggests that at least 27% of individuals infected with SARS-CoV-2 report at least one symptom three months after the acute phase [3-5], and a meta-analysis of 33 papers found that 62%, 72%, and 46% of participants reported at least one post-acute symptom at 30, 60 and 90> days respectively from infection [6]. These symptoms may continue from the acute phase or appear later on after recovery and may be persistent or occasional [7]. With such large numbers of COVID-19 cases, the burden of disease associated with post-acute consequences of COVID-19 is potentially tremendous, both at the national and global levels. Its extent and severity remain unclear.

A meta-analysis published in 2022 showed that 32% of COVID-19 patients reported fatigue or muscle weakness at 3-6 months from infection, and 41% reported the symptom after more than 12 months from infection; sleep disorders were reported by 24%, 29% and 30% of patients at 3-6, 6-9 and over 12 months respectively; and 22% reported muscle pain after more than 12 months [8]. While these symptoms, and others, are consistently reported across studies, their severity and impact on people’s well-being are not well described.

Subjective Well-Being (SWB) is the intrinsic aspect of well-being that measures the personal experience, made of the emotional and cognitive experiences [9]. The 5-item WHO Well-Being Index (WHO-5) is a commonly used and validated tool designed to measure SWB, available in more than 30 languages and in use for 24 years [10]. The WHO-5 questionnaire asks to rate on a 0-5 Likert scale five questions relating to the previous two weeks: 1. ‘I have felt cheerful and in good spirits’; 2. ‘I have felt calm and relaxed’; 3. ‘I have felt active and vigorous’; 4. ‘I woke up feeling fresh and rested’; 5. ‘My daily life has been filled with things that interest me’.

Prior to COVID-19, the effect of pandemics on SWB had not been widely studied. In a systematic review from 2015 on the use of the WHO-5 well-being index by Topp et al., none of the 213 articles included was from the field of public health [10]. This has changed slightly since the onset of the COVID-19 pandemic, with several studies measuring the effect of lockdowns and pandemic status on people’s mental health and resilience. These studies have demonstrated a detrimental effect of the pandemic on well-being across different countries and cultural contexts [11-14]. The evidence reported does not, however, measure the effect of the infection itself on SWB, let alone its post-acute symptoms.

Despite a rapidly growing body of evidence on what symptoms individuals experience following a SARS-CoV-2 infection, evidence regarding the well-being of individuals following COVID-19 and its association with reporting particular symptoms post-acute infection remains limited. We aimed to calculate the association between commonly reported symptoms following SARS-CoV-2 infection and SWB in order to better characterize the long-term impact of post-acute COVID-19 symptoms.

## Methods

### Study design and participants

This analysis uses data collected as part of a longitudinal cohort of individuals in Israel, both infected and never infected with SARS-CoV-2, who are regularly (every three-four months) asked to answer questions about their physical, mental and psychosocial health and wellbeing using a standardized questionnaire.

Details about the recruitment are described elsewhere [15]. Briefly, invitations with a link to an online survey were sent by Short Message System (SMS) to all people who had a RT-PCR SARS-CoV-2 test analyzed between July 2021 and April 2022 in one of the three governmental hospitals in Northern Israel: Baruch Padeh Poriya Medical Center, Ziv Medical Center and Galilee Medical Center. The survey was available in four languages commonly spoken in Israel: Hebrew, Arabic, Russian, and English. Participants were invited to join at three time points: July 2021, November 2021, and March 2022. Two reminders, two weeks apart, were sent to non-responders.

### Measurement tools

The study used a questionnaire designed by the International Severe Acute Respiratory and emerging Infection Consortium (ISARIC) to follow up on individuals infected with SARS-CoV-2 [16] adapted to the Israeli context. The questionnaire collected information regarding self-reported physical and mental health symptoms, quality of life, subjective well-being, daily activities, and demographics.

### Data collected

As part of the survey, participants who reported previous SARS-CoV-2 infection were asked to report symptoms they were experiencing at the time of their acute illness and in the week preceding filling the survey, from a list of 39 symptoms. Participants who reported they did not have COVID-19 were asked only about symptoms in the week preceding the survey.

All participants were also asked to answer the WHO-5 questionnaire, relating to the two previous weeks, and a numeric rating scale assessing fatigue on a 1-10 scale, regarding the last 24 hours.

### Ethics approval

The study received ethical approvals from the Ziv Medical Center (0007-21-ZIV), Baruch Padeh Medical Center (009-21-POR), and Galilee Medical Center (0018-21-NHR) ethics committees.

### Analyses

The analysis focused on participants who reported previous infection with SARS-CoV-2. SWB among individuals reporting no infection was used as a baseline for comparison. The outcome variable was the SWB score at the time of filling out the survey, consisting of five Likert scales, each with a score from 0 to 5. The raw SWB score, therefore, ranged 0-25 and was multiplied by 4 to obtain a 0-100 score.

Baseline characteristics considered in the analysis were age, sex, vaccination status (almost exclusively with the BNT162b2 vaccine), follow-up time, time of filling the survey, and symptoms reported at the time of filling the survey. Age was recorded as a continuous variable; vaccination status as binary, with anyone having received at least two doses of a COVID-19 vaccine considered vaccinated. Because of evidence of a limited association between a single dose of BNT162b2 and reporting long-term symptoms^15^, individuals who received a single vaccine dose were categorized as non-vaccinated.

Because the severity of symptoms reported post-COVID-19 is likely to change over time, participants were categorized into three groups according to the time elapsed between infection and answering the questionnaire (follow-up time): 3-6 months, 6-12 months, and 12-18 months; to avoid the conflation of acute and post-acute symptoms, patients reporting symptoms less than three months after infection were excluded. Participants reporting more than 18 months from infection were also excluded due to low numbers in this category. We also excluded participants with missing data needed for the analysis. It is important to note that participants in each of the three time periods are different. We therefore refrain from analyzing trends over time. Each of the 39 symptoms was recorded individually, and participants were asked to select the symptoms they had experienced in the week preceding filling the survey. Because of the low numbers of participants reporting rare symptoms, our analysis focused on the five most commonly reported symptoms to achieve sufficient statistical power.

Chi-square tests were performed to compare basic demographic details (sex, time survey was taken, vaccination status, and time from acute phase; student’s t-test was performed for age) between infected participants reporting experiencing and not experiencing each symptom.

For each of the three follow-up times (3-6 months post infection, 6-12 months, and 12-18 months) we used a generalized linear regression model to determine the change in SWB associated with reporting each symptom, after adjusting for the effect of other symptoms, as well as age, SARS-CoV-2 vaccination status and time of survey. We adjusted for time of survey since the survey was distributed in three waves of a month each, across three seasons and different COVID-19-related restrictions, since there is evidence that SWB is affected by seasonal changes [17]. We also present the SWB of uninfected participants, to obtain a baseline.

In addition, in order to internally validate the data, we correlated the results of the fatigue Numeric Rating Scale (NRS) included in the questionnaire (“Please rate the intensity of your fatigue on average over the last 24 hours, on a scale from 0 – 10: 0=none, 10=worst possible I can imagine.”) with mean SWB scores of participants using Pearson correlation tests, and performed a student’s t-test to compare fatigue scale scores between participants reporting and not reporting experiencing fatigue in the symptoms questionnaire.

Data were collected online using the Alchemer online survey solution (Louisville, CO), cleaned using Microsoft Excel and analyzed using R studio (Boston, MA).

## Results

Of 95604 people invited, 6500 answered the survey (6.8%). After excluding those with incomplete data and those not meeting the eligibility criteria, 2295 participants were included in the analysis (Figure 1).

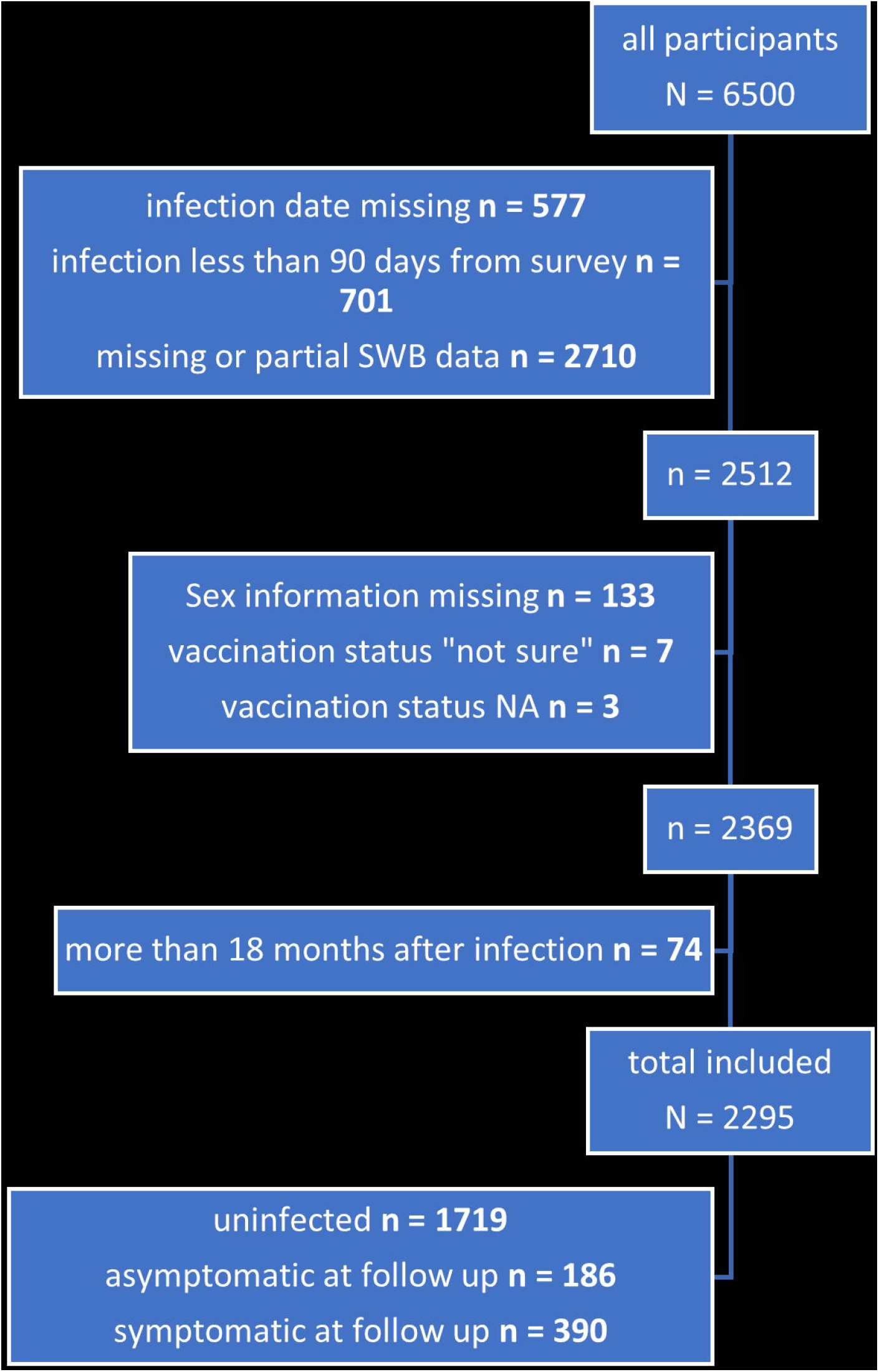

Overall, 1319 of the 2295 participants (57%) were female, 2112 (92%) were vaccinated, and 1719 (75%) reported no previous infection. Of the 576 who reported previous infection, 186 (32%) did not report any symptoms at time of survey, and 390 (68%) reported experiencing at least one symptom. The median time between infection and responding to the survey was 260 days (270 days among those reporting any symptoms and 256 days among those not reporting symptoms). Those reporting symptoms post infection were more likely to be female, and vaccinated individuals reported fewer symptoms than those unvaccinated, as previously reported [15,19]. Baseline characteristics of those reporting and not reporting symptoms were otherwise similar (Table 1).

**Table 1:** Demographics and chi-square tests of cohort participants, infected only (N=576), 3-18 months from infection.

|  | Fatigue |  |  | Weakness in muscles |  |  | Muscle pain |  |  | Lack of concentration |  |  | Sleep disruption |  |  |
| --- | --- | --- | --- | --- | --- | --- | --- | --- | --- | --- | --- | --- | --- | --- | --- |
|  | No<br>(N=333) | Yes<br>(N=243) | p-value | No<br>(N=423) | Yes<br>(N=153) | p-value | No<br>(N=442) | Yes<br>(N=134) | p-value | No<br>(N=450) | Yes<br>(N=126) | p-value | No<br>(N=455) | Yes<br>(N=121) | p-value |
| Time from acute phase |  |  |  |  |  |  |  |  |  |  |  |  |  |  |  |
| 3-6 months | 129<br>(62%) | 79 (38%) | 0.133 | 164<br>(78.8%) | 44 (21.2%) | 0.082 | 165<br>(79.3%) | 43 (20.7%) | 0.539 | 169<br>(81.2%) | 39 (18.8%) | 0.357 | 169<br>(81.2%) | 39 (18.8%) | 0.306 |
| 6-12 months | 107<br>(52.5%) | 97 (47.5%) |  | 145<br>(71.1%) | 59 (28.9%) |  | 154<br>(75.5%) | 50 (24.5%) |  | 154<br>(75.5%) | 50 (24.5%) |  | 154<br>(75.5%) | 50 (24.5%) |  |
| 12-18 months | 97<br>(59.1%) | 67 (40.9%) |  | 114<br>(69.5%) | 50 (30.5%) |  | 123<br>(75%) | 41 (25%) |  | 127<br>(77.4%) | 37 (22.6%) |  | 132<br>(80.5%) | 32 (19.5%) |  |
| Sex |  |  |  |  |  |  |  |  |  |  |  |  |  |  |  |
| female | 200<br>(52.5%) | 181 (47.5%) | <.001 | 267<br>(70.1%) | 114 (29.9%) | 0.014 | 277<br>(72.7%) | 104 (27.3%) | 0.002 | 284<br>(74.5%) | 97 (25.5%) | 0.005 | 301<br>(79%) | 80 (21%) | 1 |
| male | 133<br>(68.2%) | 62 (31.8%) |  | 156<br>(80%) | 39 (20%) |  | 165<br>(84.6%) | 30 (15.4%) |  | 166<br>(85.1%) | 29 (14.9%) |  | 154<br>(79%) | 41 (21%) |  |
| ARS-CoV-2 vaccination (at least 2 doses) |  |  |  |  |  |  |  |  |  |  |  |  |  |  |  |
| No | 174<br>(54.5%) | 145 (45.5%) | 0.092 | 220<br>(69%) | 99 (31%) | 0.009 | 238<br>(74.6%) | 81 (25.4%) | 0.212 | 241<br>(75.5%) | 78 (24.5%) | 0.118 | 241<br>(75.5%) | 78 (24.5%) | 0.031 |
| Yes | 159<br>(61.9%) | 98 (38.1%) |  | 203<br>(79%) | 54 (21%) |  | 204<br>(79.4%) | 53 (20.6%) |  | 209<br>(81.3%) | 48 (18.7%) |  | 214<br>(83.3%) | 43 (16.7%) |  |
| Age |  |  |  |  |  |  |  |  |  |  |  |  |  |  |  |
| Mean (SD) | 46.9<br>(15.1) | 44.0 (13.4) | 0.013 | 45.9<br>(14.8) | 44.9 (13.6) | 0.461 | 45.7<br>(14.8) | 45.7 (13.2) | 0.952 | 46.2<br>(14.8) | 44.0 (13.1) | 0.14 | 45.5<br>(14.7) | 46.5 (13.6) | 0.481 |
| Median<br>[Min, Max] | 45<br>[18, 94] | 41 [19, 88] |  | 44<br>[18, 94] | 43 [19,88] |  | 43<br>[18,94] | 44 [24,88] |  | 44<br>[18, 94] | 42 [21, 88] |  | 43<br>[18,94] | 46 [20,88] |  |
| Time of survey (season and year) |  |  |  |  |  |  |  |  |  |  |  |  |  |  |  |
| Summer 21' | 57<br>(49.1%) | 59 (50.9%) | 0.106 | 79<br>(68.1%) | 37 (31.9%) | 0.315 | 86<br>(74.1%) | 30 (25.9%) | 0.469 | 89<br>(76.7%) | 27 (23.3%) | 0.567 | 84<br>(72.4%) | 32 (27.6%) | 0.029 |
| Winter 21' | 152<br>(59.8%) | 102 (40.2%) |  | 192<br>(75.6%) | 62 (24.4%) |  | 201<br>(79.1%) | 53 (20.9%) |  | 195<br>(76.8%) | 59 (23.2%) |  | 197<br>(77.6%) | 57 (22.4%) |  |
| Spring 22' | 124<br>(60.2%) | 82 (39.8%) |  | 152<br>(73.8%) | 54 (26.2%) |  | 155<br>(75.2%) | 51 (24.8%) |  | 166<br>(80.6%) | 40 (19.4%) |  | 174<br>(84.5%) | 32 (15.5%) |  |

At the time of responding to the survey, among participants included in the analyses who report having COVID-19 (n= 576) the five most common symptoms were: fatigue (n = 243, 42%), weakness in muscles (n = 153, 26%), muscle pain (n = 134, 23%), confusion/ lack of concentration (n = 126, 22%), and sleep disorders (n = 121, 21%).

The overall SWB score of those previously infected but reporting no symptoms post-infection was similar to those reporting never having been infected (74.3, 95%CI 71.3-76.7 vs. 73.7, 95%CI 72.7-74.8). Among commonly reported symptoms, sleeping disorders, lack of concentration, and fatigue were each independently associated with a significant overall decrease in SWB of 11.5, 10.7, and 7.7 percentage points, respectively (p< .002 for all, Table 2), as well as muscle pain with 6.5 percentage points significant decrease (p = .05).

**Table 2:**
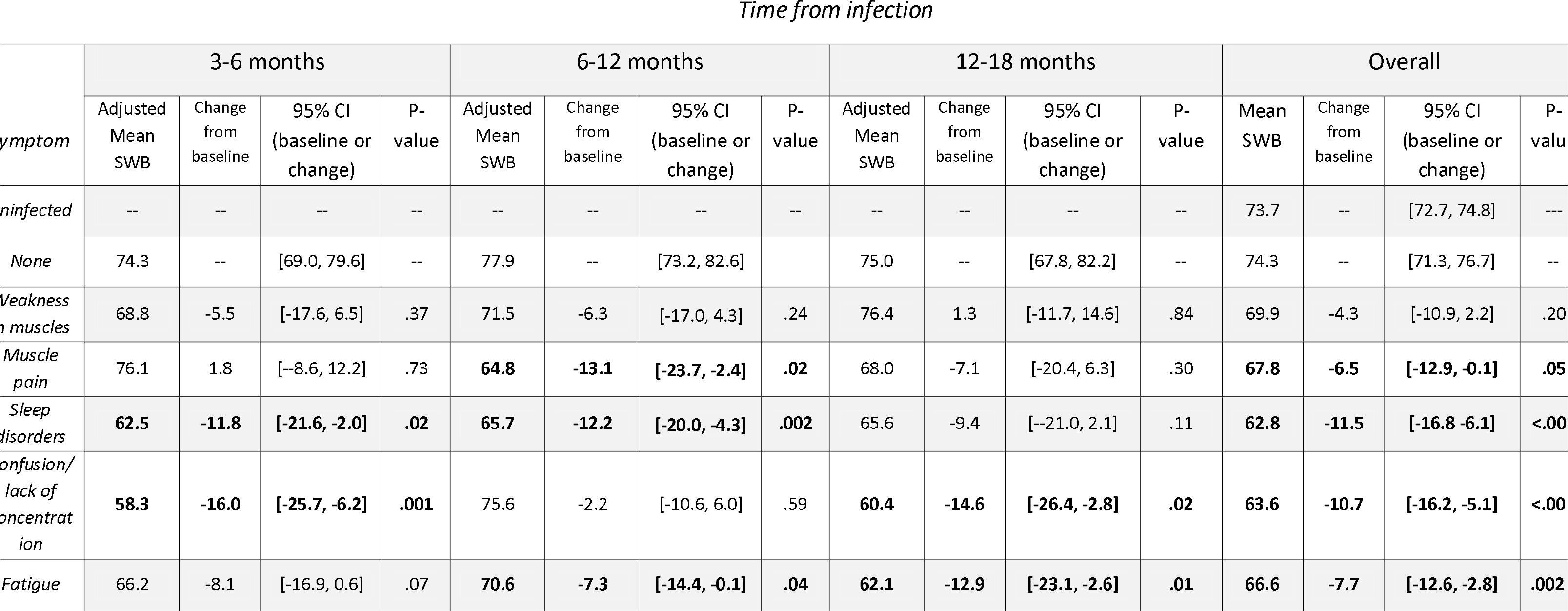
Regression model results for adjusted mean SWB per reported symptom, change from no reported symptoms, 95% CI and p-value. adjusted over age, vaccination status, other symptoms reported (yes/no) and time of survey.

| *ymptom* | 3-6 months |  |  |  | 6-12 months |  |  |  | 12-18 months |  |  |  | Overall |  |  |  |
| --- | --- | --- | --- | --- | --- | --- | --- | --- | --- | --- | --- | --- | --- | --- | --- | --- |
|  | Adjusted Mean SWB | Change from baseline | 95% CI (baseline or change) | P-value | Adjusted Mean SWB | Change from baseline | 95% CI (baseline or change) | P-value | Adjusted Mean SWB | Change from baseline | 95% CI (baseline or change) | P-value | Mean SWB | Change from baseline | 95% CI (baseline or change) | P-value |
| *ninfected* | -- | -- | -- | -- | -- | -- | -- | -- | -- | -- | -- | -- | 73.7 | -- | [72.7, 74.8] | --- |
| *None* | 74.3 | -- | [69.0, 79.6] | -- | 77.9 | -- | [73.2, 82.6] |  | 75.0 | -- | [67.8, 82.2] | -- | 74.3 | -- | [71.3, 76.7] | -- |
| *Weakness in muscles* | 68.8 | -5.5 | [-17.6, 6.5] | .37 | 71.5 | -6.3 | [-17.0, 4.3] | .24 | 76.4 | 1.3 | [-11.7, 14.6] | .84 | 69.9 | -4.3 | [-10.9, 2.2] | .20 |
| *Muscle pain* | 76.1 | 1.8 | [-8.6, 12.2] | .73 | **64.8** | **-13.1** | **[-23.7, -2.4]** | **.02** | 68.0 | -7.1 | [-20.4, 6.3] | .30 | **67.8** | **-6.5** | **[-12.9, -0.1]** | **.05** |
| *Sleep disorders* | **62.5** | **-11.8** | **[-21.6, -2.0]** | **.02** | **65.7** | **-12.2** | **[-20.0, -4.3]** | **.002** | 65.6 | -9.4 | [-21.0, 2.1] | .11 | **62.8** | **-11.5** | **[-16.8 -6.1]** | **<.00** |
| *Confusion/ lack of concentration* | **58.3** | **-16.0** | **[-25.7, -6.2]** | **.001** | 75.6 | -2.2 | [-10.6, 6.0] | .59 | **60.4** | **-14.6** | **[-26.4, -2.8]** | **.02** | **63.6** | **-10.7** | **[-16.2, -5.1]** | **<.00** |
| *Fatigue* | 66.2 | -8.1 | [-16.9, 0.6] | .07 | **70.6** | **-7.3** | **[-14.4, -0.1]** | **.04** | **62.1** | **-12.9** | **[-23.1, -2.6]** | **.01** | **66.6** | **-7.7** | **[-12.6, -2.8]** | **.002** |

The impact of individual symptoms on SWB was different according to time point. Whereas fatigue led to a non-significant 8.1 percentage points decrease in SWB in those reporting the symptom at 3-6 months post infection, among those reporting it at 12-18 months, the decrease in SWB associated with fatigue was larger and statistically significant (−12.6, p=0.01 respectively). Lack of concentration was associated with a large decrease in SWB both when reported 3-6 months post COVID-19 (−16.0, p = 0.001) and 12-18 months (−14.6, p = .02). The impact of sleep disorders on SWB was significant at 3-6 months and 6-12 months post COVID-19 (−11.8, p = .02; −12.1, p = .002, respectively), but not after 12-18 months. Physical musculoskeletal symptoms (muscle weakness and muscle pain) were not significantly associated at any of the follow-up times except for a decrease in SWB among those reporting muscle pain 6-12 months post infection (−13.1, p = .02), (Figure 2).

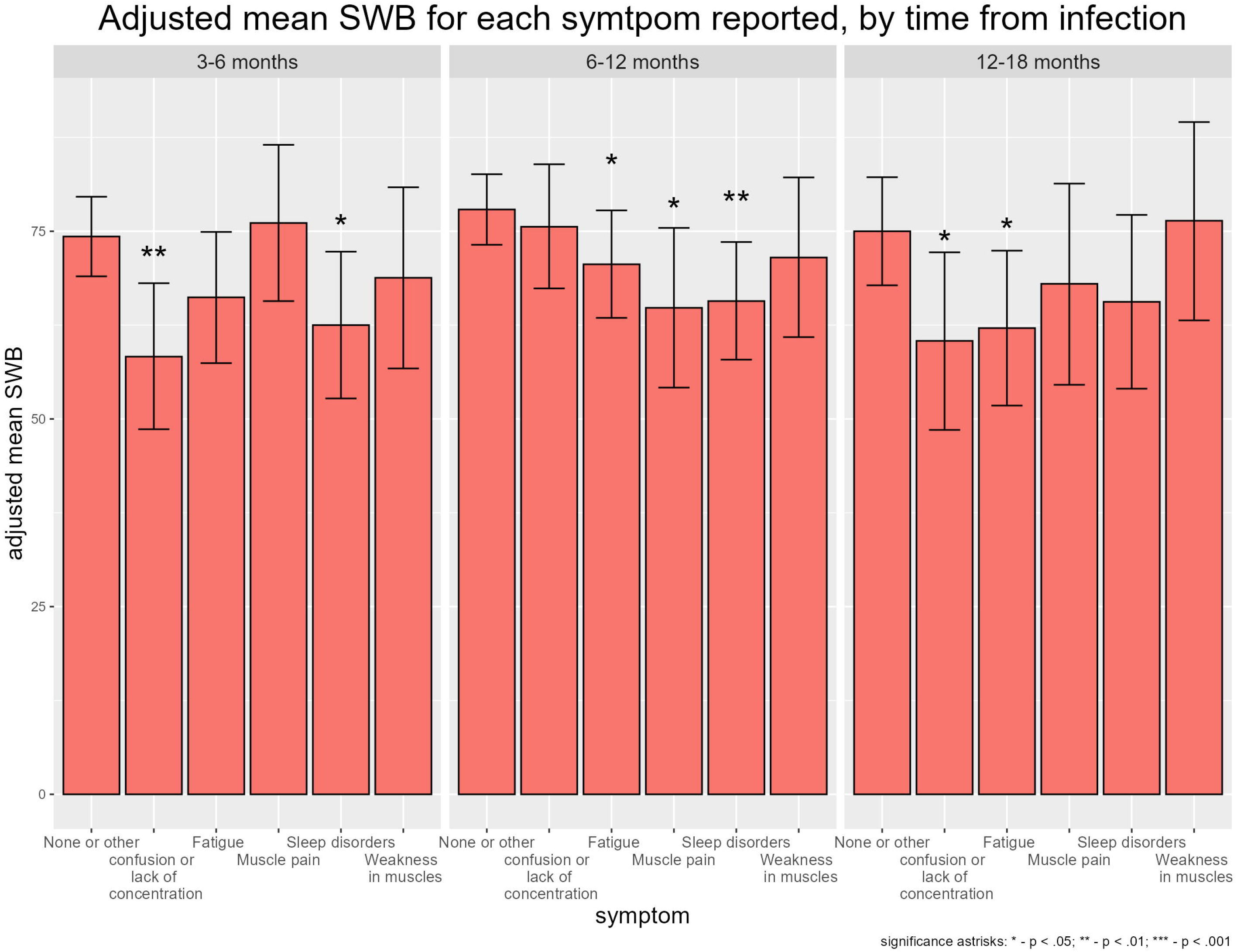

Among the 553 infected participants who completed the NRS, participants reporting fatigue had a significantly higher NRS score (more fatigue) than those who did not (7.06 vs. 4.13, p <.001). SWB decreased with increasing NRS score (r = −0.54, 95% CI: [−0.60, −0.48], p<.001).

## Discussion

There is an increasing amount of evidence that a substantial proportion of patients experience a wide range of symptoms following infection with SARS-CoV-2. Few studies, however have attempted to quantify their impact on wellbeing. This is, to our knowledge, the first study to disentangle the effect on the SWB of the most commonly reported symptoms post COVID-19.

Our analysis shows that not all symptoms impact patients in the same way, or necessarily consistently over time. Compared with physical musculoskeletal symptoms, symptoms that are more nebulous and harder to attribute to specific physiological systems such as fatigue, confusion/ lack of concentration, and sleeping difficulties, have a more profound and more sustained impact on SWB. Whereas the impact on SWB of sleep disorders, fatigue, or lack of concentration was similar or larger in those reporting these symptoms 12-18 months post infection than in those reporting them 3-6 months post infection, the strongest decrease in SWB for musculoskeletal symptoms was seen at 6-12 months post infection, with no significant change in SWB after that. This could be interpreted as these symptoms becoming less severe over time, or individuals learning to cope with persisting physical symptoms. Such as phenomenon has been documented for other communicable diseases post infection [18]. By contrast, the lack of change in associations between SWB and fatigue, confusion/ lack of concentration, and sleeping difficulties could suggest the cumulative impact of these symptoms, no improvement in these symptoms over time, or more difficulty in adapting to these symptoms. This study is not a cohort study, and each time point represents different individuals – it is therefore challenging to comment on trends over time.

In this study, the association between SWB and each symptom is adjusted for reporting other symptoms. This implies that, in particular, fatigue, lack of concentration, and sleep disorders were independently associated with large changes in SWB. Because a substantial proportion of patients report, more than one of these symptoms, the cumulative effect of symptoms reported post COVID-19 on SWB may be profound.

Several elements of the analysis validate the internal consistency of our data: first, the similar SWB scores between those never infected and those reporting no post-COVID-19 symptoms; second, the association between a high score on the NRS and fatigue reporting, and decreasing SWB with increasing NRS scores. This validates using SWB as an indirect measurement of the impact of specific symptoms, at least for fatigue.

Although those reporting and not reporting symptoms were comparable in terms of baseline characteristics, females reported having more of most symptoms than males. This is consistent with other studies showing females are more likely to report post-acute COVID-19 symptoms than men [19,20].

In Israel, the WHO-5 questionnaire has not been used extensively and lacks a formal translation despite the large amount of health quality-related studies in Israel. The few studies found focus mainly on minority populations such as immigrant creworkers [21] or Bedouin caregivers [22]. We found only one study that surveyed the WHO-5 in the general population [23], and found an average of 55.6 points (SD = 24.8). This is lower than in our study and lower than most countries reported in the same paper in Europe (e.g. Germany-63.59, United Kingdom – 62.51, Netherlands – 65.29), and in other Mediterranean countries such as Cyprus (58.31), Greece (60.37), Italy (58.31) and Turkey (58.43) [24]. Such comparisons are possible since SWB in different countries can be compared since the WHO-5 scale was found to take transcultural components into consideration [25].

In our study, the mean SWB of all participants in the survey (n = 3113) was 69.6 (SD = 24.4), closer to Northern European countries than other Mediterranean ones. A high SWB in the Israeli population is plausible since Israel regularly reports high levels of happiness in the population and ranked 9^th^ in the World happiness report 2022 [26]. The difference could also be explained by the fact that our sample was not designed to be representative of the general population. In our samples, females are overrepresented and the very elderly are underrepresented.

There are some limitations to this study. The first limitation, shared by all studies using questionnaires for self-reported health status, is that we cannot infer whether symptoms reported after the acute episode are directly attributable to COVID-19 or not. In the absence of a case definition specifying which post-acute symptoms can be attributed to COVID-19 infection, our approach preferred sensitivity to specificity and included all reported symptoms.

A second limitation is the cross-sectional nature of the study. While the questionnaire design ensures that SARS-CoV-2 infection predates the reporting of symptoms, participants are different at each time point. This potentially introduces selection bias since individuals with severe post-acute symptoms may be more interested in participating. The study invitation mentioned health in general during the COVID-19 pandemic and did not specifically mention a focus on post-acute symptoms, thus reducing this possibility.

Another limitation is that our study does not measure the duration or severity of reported symptoms. Subjective wellbeing is a complex phenomenon, and changes over time can have alternative explanations other than changes in severity. For example, normalizing symptoms that do not change in severity can lead to an increase in SWB. As a result, SWB should not be interpreted as a marker of severity, but rather of impact. However, the association between changes in SWB and NRS suggests that, at least for fatigue, using SWB as a proxy for severity may be reasonable. Future studies using our cohort will make use of several data points per participant and will help quantify the duration of symptoms.

Our cohort largely represents individuals who suffered mild to moderate COVID-19. Since the severity of post-acute symptoms correlates with the severity of the initial acute episode [27], our findings may not be applicable to individuals who suffered severe initial disease.

Our study is, to our knowledge, the first to show a substantial and sustained impact of post-acute symptoms on well-being following infection with SARS-CoV-2. At 12-18 months following acute infection, a decrease in SWB was still reported, particularly associated with non-specific symptoms commonly reported post-COVID-19, such as fatigue and lack of concentration. Because reporting more than one of these symptoms is common, the cumulative impact of such symptoms on SWB can be profound. Understanding the underlying biological causative mechanisms of these long-term symptoms will be central to developing treatment to mitigate what could become a serious public health issue post pandemic. We plan to follow individuals recruited into our cohort over time to better understand trends of reported symptoms over time, their duration, and their changing impact on well-being and quality of life.

## Data Availability

The dataset used in this study is available upon request to the corresponding author

## Acknowledgments

We wish to thank Mark Lifshitz and Shelly Shalem of Galilee Medical Center and Yehudit Hachmon and Eliran Levi of Ziv Medical center for their help in data collection, management, and general support.

## Data availability statement

the dataset used in this analysis is available upon request to the corresponding author

